# Exploring the Decisional Needs of Patients living with Subacromial Pain Syndrome: a qualitative needs assessment study

**DOI:** 10.1101/2024.10.09.24314833

**Authors:** Samantha Charmaine Bengtsen, Joshua Robert Zadro, Michael Skovdal Rathleff, Nadine E Foster, Janus Laust Thomsen, Jens Lykkegaard Olesen, Jens Søndergaard, Kristian Damgaard Lyng

**Author notes:** **Contact information for the corresponding author** Kristian Damgaard Lyng, Department of Health Science and Technology, Aalborg University Selma Lagerlöfs Vej 249, 9260 Gistrup, Denmark.

## Abstract

**Background:** There are a variety of different treatments for patients living with subacromial pain syndrome (SAPS). All treatments have small to moderate effect sizes, and it is challenging when healthcare practitioners and patients need to decide on which treatment options to choose. The aim of this study was to explore and understand the decisional needs of patients with SAPS, to inform and support the decision-making process.

**Methods:** A qualitative research study, using semi-structured individual interviews with patients with SAPS. The interview guide was informed by the Ottawa Decision Support Framework (ODSF), previous research related to treatment decision-making, other decisional needs assessment studies, and inputs from patients with SAPS and healthcare practitioners. Data were analysed by using reflexive thematic text analysis and ODSF. The analysis was conducted in NVivo 12.

**Results:** We invited 22 participants of which 17 (age 22-71 years) took part in the study. We found three main themes related to individual decisional needs in the context of decision-making: 1) The necessity of certainty and adequate information as fundamental prerequisites for effective decision-making, 2) The importance of person-centred care to achieve a desirable decision, and 3) The need for a supportive environment to facilitate adaptation and acceptance of the decision.

**Conclusion:** The decision-making process faced by patients with SAPS is complex and involves several decisional needs. Our findings highlight the importance of healthcare professionals identifying and addressing patients’ decisional needs in consultations with patients with SAPS.

**Implications:** Our study provides guidance to clinical practice in terms of increased understanding of the decisional needs experienced by patients with SAPS. Furthermore, these results should inform future development of decision aids for patients with SAPS.

**Highlights:**

- Subacromial pain syndrome is a complex condition, demanding a thorough decision-making process
- We identified several concrete decisional needs through the Ottawa Decision Support Framework and further provided in-depth knowledge on key themes related to certainty, patient-centeredness, and support.
- Our study provides information of designing a decision aid supporting shared decision-making in future studies.

## Introduction

Subacromial pain syndrome (SAPS) is a common musculoskeletal condition, characterised by pain and functional impairment in the shoulder region [1]. Treatments for patients living with SAPS include pain medications (e.g. paracetamol or nonsteroidal anti-inflammatory drugs), exercise (e.g. resistance exercises), injections (e.g. corticosteroid), or surgical interventions (e.g., subacromial decompression) [1]. These treatments are frequently provided individually and in combination [1,2]. There is uncertainty about whether some types of treatment are superior. Research evidence shows small to moderate effect sizes when these treatment options are compared, which makes the decision-making process challenging and preference-sensitive [1–3]. As a result, patients with SAPS who seek care need to consider the trade-offs between the benefits and harms of two or more broadly equivalent treatment options [4]. Our previous research showed that by identifying key treatment attributes valued by patients, healthcare providers may be more able to effectively individualise care to an individual’s needs [5]. Failure to adequately identify and address patients’ needs during the clinical consultation may lead to decisional conflicts, and even treatment regret among some patients [6]. This can occur due to a clinician-driven paternalistic approach to the decision-making process, undermining the patient’s autonomy and sense of certainty in relation to their conditional state [7,8]. Shared decision-making (SDM) aims to bridge all stakeholder’s needs and preferences in the decision-making process and is commonly seen as an approach to accommodate preference-sensitive decisions [4,9]. Recent studies have shown how SDM improves communication between stakeholders and condition specific knowledge in patients with chronic pain, leading to better health outcomes, including reduced pain [10–12]. However, applying SDM in the context of decision-making and patients living with SAPS requires a deeper understanding of the individual’s decisional needs, including how psychosocial, emotional, and contextual factors influence decision-making. The overall aim of this study was to explore and understand the decisional needs of patients living with SAPS.

## Methods

### Design

We conducted a descriptive qualitative study using semi-structured, individual interviews with patients with SAPS. The reporting follows the Standards for Reporting Qualitative Research (SRQR) [13]. The study was guided by the Ottawa Decision Support Framework (ODSF), chosen to explore and understand the decisional needs of PwSAPS, making it suitable for informing a future decision aid for this patient group (See **Supplementary File S1: ODSF**) [8]. The ODSF aims to guide clinicians in assessing the decisional needs of patients with lived experience of a specific health condition and to support clinicians in providing patients with decisional support [7,8]. Furthermore, the ODSF was developed to facilitate researchers in identifying decisional needs and evaluating decisional outcomes such as decisional conflict and regret [7,8]. The ODSF is centered around three factors: decisional needs, decision support, and decisional outcomes. This study formed the initial step in a research programme developing a patient decision aid for PwSAPS. The full protocol for the research programme, including this study, was preregistered on Open Science Framework [14].

### Researcher characteristics and reflexivity

Our interprofessional research group consisted of experienced clinician-researchers (physiotherapists, rheumatologists, and general practitioners) within the field of musculoskeletal pain, including those with experience conducting research in SAPS. The lead investigators (SCB and KDL) are both physiotherapists. SCB has seven years of clinical experience with patients with SAPS, and KDL is a PhD candidate with seven years of experience in musculoskeletal pain research and qualitative research. The other members of the study team (JRZ, JLO, MSR, NEF, JLT, and JS) are experienced researchers with a combined publication record of over 1,200 peer-reviewed articles and extensive expertise in musculoskeletal pain and qualitative research. We acknowledge that the results presented in this paper will be influenced by the teams’ preconceptions and perspectives.

### Study population

We included patients aged 18 years and over, previously diagnosed with SAPS (or a related diagnosis e.g. rotator cuff related shoulder pain) by their healthcare practitioner. Patients were eligible if they had had at least one clinical consultation with an authorised healthcare practitioner and had been involved in at least one treatment decision-making situation. Patients were excluded if their pain had a traumatic onset, or if they had any neurological symptoms or other concomitant diagnoses interfering with their ability to participate or possibly contaminate their answers relevant to the aim of this study. Furthermore, patients were excluded if they had any cognitive or communicative challenges or were unable to speak or understand Danish. We anticipated needing to interview up to 20 participants, and our sample size was guided by data saturation (i.e., when saturation was reached in terms of key themes, we conducted two more interviews to ensure no new data emerged).

### Sampling strategy

Patients were identified using several methods, including identification from physiotherapy and general practice clinics in the Capital City Region, Mid-Jutland Region, and North Jutland Region of Denmark. Furthermore, we included patients from our previous research studies who had given consent to be contacted about future research, and through posts on a shoulder pain dedicated channel on Facebook and Instagram. Patients identified from clinics were informed by their clinicians about the study and in cases of interest, patients provided their contact information and were then contacted by the study team who then screened patients for eligibility. Patients were screened over the phone and eligible patients were informed about the study by one of two authors (SCB or KDL).

### Data Collection

A semi-structured interview guide was developed based on the OSDF, scientific literature in relation to SDM, decision aids and needs assessments, and discussions with stakeholders (e.g., patients living with SAPS, clinicians, and researchers) from the larger research programme (**Supplementary File S2**). Each interview participant was asked a short series of questions about their age, gender, duration of pain, highest level of education, previous shoulder treatments and consultations with healthcare practitioners. The core part of the interview guide asked participants about their understanding of their condition/diagnosis, their thoughts and experiences of the decision-making process related to their choice of treatment, how their values and preferences matched their treatment choice, how the decision-making process was aided and lastly, how potential uncertainties about treatment options were managed. The interview guide consisted of multiple probes to explore further, in line with the semi-structured interview approach. All interviews were conducted over the telephone after at least one week of commencing treatment (as self-reported by each participant). The principal investigator (KDL) and the research coordinator (SCB) conducted all interviews, and all interviews were audio-recorded with consent. Data on participant characteristics were collected and managed using the secure, web-based software REDCap (Research Electronic Data Capture) hosted at Aalborg University [15,16]. All audio-recordings were saved on a secure server hosted at Aalborg University to adhere to the existing EU-GDPR and Aalborg University guidelines for storing data. All participant’s names were noted in an encrypted file that was stored in a separate folder.

### Research Ethics

Written informed consent (via a digital consent form forwarded to each patient) and oral informed consent was obtained from all participants before the interviews. Findings are presented anonymously, adhering to research ethical standards, ensuring the protection of participants’ identity. The study was conducted in accordance with the Helsinki Declaration and was deemed exempt from full ethical approval by The North Denmark Region Committee on Health Research Ethics (2023-000206).

### Data Analysis

All audio-recorded interviews underwent verbatim transcription independently by two researchers (SCB and KDL) and details of names and recognisable places were removed. All transcripts were imported to NVivo (QSR International Pty Ltd, version 11.4, United Kingdom) for analysis. The analysis was performed independently by SCB and KDL using the reflexive approach to thematic text analysis, as previously described by Braun and Clarke [17]. The analysis was initiated by a naïve readthrough and familiarisation of the transcripts to attain an overview and overall understanding of the texts. Initially, a set of codes was created and thereafter, proposed to the study team. The initial set of codes was discussed at the preliminary review stage. Subsequent coding rounds were carried out to identify additional new codes, with discussions and documentation within the coding list occurring progressively throughout the analysis. Through inductive grouping and merging of the codes, main themes were identified reflexively, centered around the descriptions from the patients living with SAPS and their decisional needs. From here, a hierarchical mind-map was developed to organise the main themes and sub-themes, which were then summarised into an overall description of the narrative. Furthermore, the codes were deductively visualised using ODSF to conceptualise and formalise the decisional needs, relevant to the aim of the study, using direct quotes from the interviews. Coding followed the coding manual formulated by Hoefel et al. (**Supplementary File S3**) [7,18]. Supporting quotes were translated to English for the purpose of this paper.

## Results

Of 22 potentially eligible patients, 20 were deemed eligible and 17 provided consent and were interviewed and included in the analysis. Two patients were deemed ineligible because of other concomitant diagnoses and three due to their lack of willingness to participate, and two because of illness. Data saturation was judged to be reached after 15 interviews, and then two additional interviews were conducted, and no additional themes were identified. Participants’ characteristics are summarised in **Table 1**. We identified three main themes relevant to patients’ decisional needs, *Certainty and Information as Prerequisites for Decision-making*, *Person-centered care as a Need for a Desirable Decision*, *and Supportive Environment as a Need for Adapting and Accepting the Treatment Plan*. An overview of the themes and their connections are visualised in **Figure 1**. Furthermore, the findings were mapped to the OSDF, which can be seen in **Table 2**. A detailed description of all themes is presented below.

**Figure 1:**
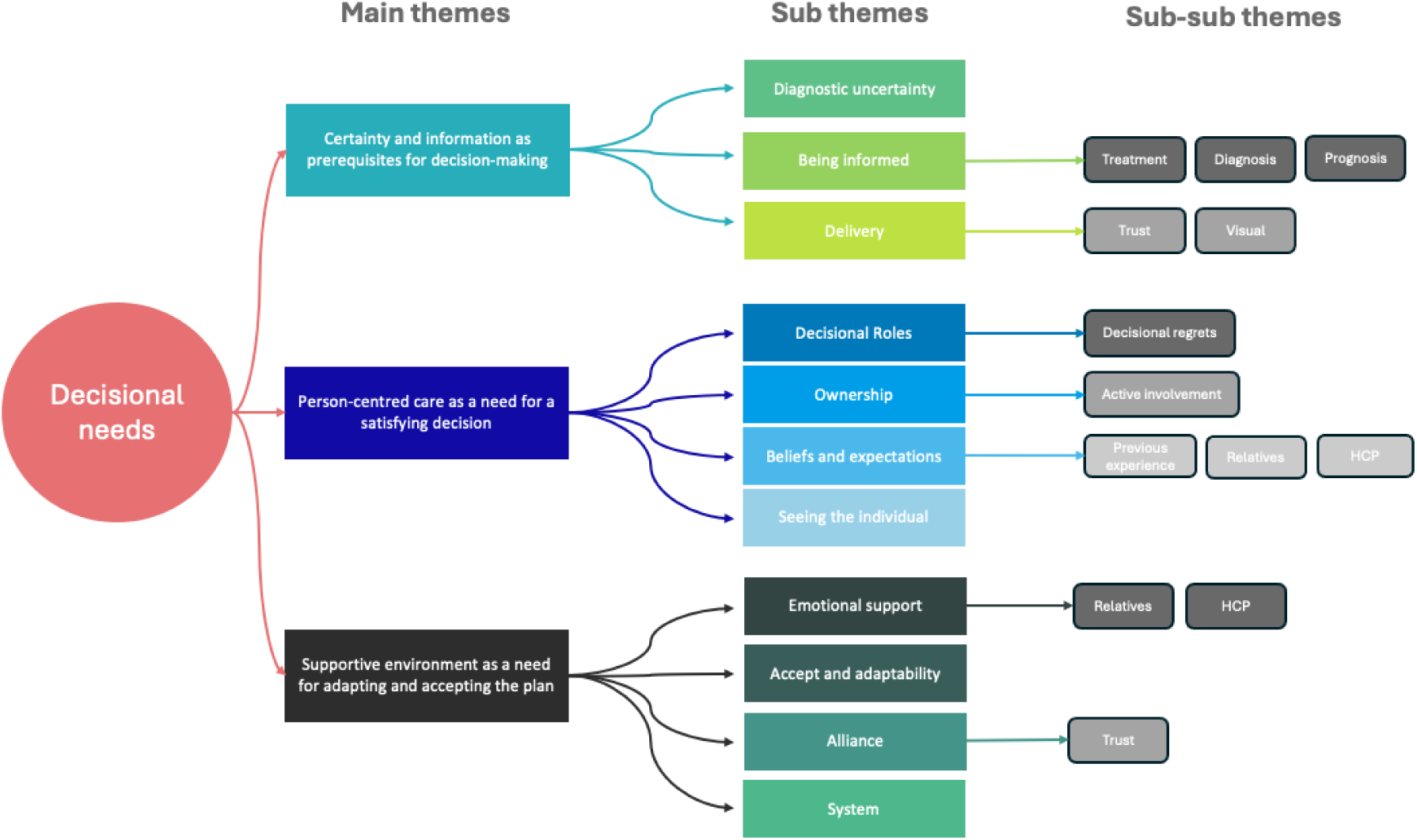
Overview of main themes, sub-themes, and sub-sub themes. HCP = Healthcare practitioner.

**Table 1:** Interview participants characteristics.

| **Table 1. Interview participants characteristics (n=17)** |  |  |
| --- | --- | --- |
| **Female, n (%)** | 10 (59%) |  |
| **Age, median in years (range)** | 49 (22-71) |  |
| **Duration of pain, months (range)** | 24 (3-120) |  |
| **Education (ISCED\*), n (%)** | **n, %** |  |
|  | Upper secondary education | 5 (29%) |
|  | Short-cycle tertiary education | 4 (23.5%) |
|  | Bachelor's or equivalent | 3 (18%) |
|  | Master's or equivalent | 4 (23.5%) |
|  | Doctorate or equivalent | 1 (6%) |
| **Treatments previously received, n (%)** | **n, %** |  |
|  | Pain medication | 17 (100%) |
|  | Exercise | 16 (94%) |
|  | Education | 15 (88%) |
|  | Injection | 11 (64%) |
|  | Manual therapy | 10 (59%) |
|  | Shockwave | 7 (41%) |
|  | Surgery | 6 (35%) |
|  | Other | 7 (41%) |
| **Healthcare practitioners previously consulted, n (%)** | **n, %** |  |
|  | General Practitioner | 17 (100%) |
|  | Physiotherapist | 15 (88%) |
|  | Orthopedic Surgeon | 11 (64%) |
|  | Chiropractor | 7 (41%) |
|  | Rheumatologist | 4 (23.5%) |
|  | Psychologist | 2 (12%) |
|  | Other (examples included Osteopaths and Complementary Medicine) | 8 (47%) |
| **Most recent patient consultations with healthcare practitioner, n (%)** | **n, %** |  |
|  | General Practitioner | 9 (52%) |
|  | Physiotherapist | 7 (41%) |
|  | Orthopedic Surgeon | 1 (6%) |
|  | Rheumatologist | 0 (0%) |
|  | Chiropractor | 0 (0%) |
|  | Psychologist | 0 (0%) |
|  | Other | 0 (0%) |
|  | **n, %** |  |
| **Recruitment, n (%)** | Physiotherapy | 5 (29%) |
|  | Social media | 5 (29%) |
|  | Previous research studies | 4 (23.5%) |
|  | General practitioner | 3 (18%) |

\* The International Standard Classification of Education

### Certainty and Information as Prerequisites for Decision-making

This theme explored the importance of how appropriate and sufficient information, coupled with a sense of certainty, were key drivers for limiting decisional regret and conflict in the decision-making process. Furthermore, participants expressed their preferred information format, the need for knowledge on prognosis and the potential costs of various treatments. Overall, participants expressed that the most important prerequisite for a satisfactory decision-making process was that the individual was generally informed about existing treatment options, their effectiveness, and their risks and benefits.

> **ID10**: “*Well, I did have a need for more information on why I was asked to say yes to that (exercise) option. I generally don’t really feel like I know why I was doing it; I just trusted my gut and my doctor and physiotherapist. In retrospect, I’m sure that I would have picked differently if I had the feeling that I was more informed”*.

This informational need also included obtaining knowledge on their condition “*label”* (**ID5**) or diagnosis as well as on prognosis and impact on workability. Several participants emphasised how the lack of an early and clear label or diagnosis for their shoulder problem, hampered their trust in their healthcare practitioner and delayed them commencing their treatment plan, leaving them unsatisfied and frustrated with their care.

> **ID9**: *“My doctor was unsure what my diagnosis was, so I was waiting on him to refer me to a physio, the physio didn’t want to touch my shoulder before she had a scan of my shoulder, which was fair. However, I waited four months before I got the message that it was nothing dangerous and we could just start exercise – why couldn’t I have done anything in the meantime?”*.

It was further stressed that in some more severe cases, if the need for a diagnosis wasn’t met, this led to a sense of feeling stigmatised.

> **ID2**: *“I was circulated back and forth for years in the (healthcare) system. Nobody could figure out what was the cause of my pain. At some point, I remember that a surgeon asked me if this was something I made up in my head. I will never forget that, but sadly, it is not the only time that I have had people being sceptical about my pain and not believing me – that’s why partly also why I have been searching so much for someone who can give me a diagnosis”*.

Two participants expressed the need for more information in terms of the potential costs of treatments, especially in the longer-term. One of the participants expressed unsatisfactory experiences, because they were unaware that they needed to pay for certain aspects of the treatment themselves.

> **ID11**: “*Looking back on my care, I would have appreciated more knowledge on how much injections would have cost me over the years to keep this (the pain) in check. If I had known how much that treatment would have cost me, I would rather have been doing a few exercises now and then”*.

All participants were willing to get their information through the internet, however their preferred way of obtaining new knowledge, was together with their general practitioner, or other healthcare practitioners, where they were able to ask questions and get answers.

> **ID2**: “*I don’t care which clinician is giving me the information (on treatment options), but I needed it from a health-person whom I can trust and talk through the options. I had already read a lot on the internet, before my first consultation, but I did have some questions that the internet couldn’t answer”*.

Three participants saw the internet as a barrier for acquiring new knowledge, as they were often confused by conflicting information on different websites. Importantly, several participants noted that the information had to be provided by a reliable source, hence their preferred information source was either through governmental sites (e.g., Sundhed.dk) or through their healthcare practitioner.

> **ID6**: “*I think that the information I receive should be given balanced and unbiased. Sometimes I’m afraid that what I read on different websites is conflicting and maybe it is because they have different interests in selling me something. So, I try to keep away from the internet”*.

Of the 17 participants, 14 expressed the need for a visual product that could facilitate their learning of information about their condition. In this regard, several participants expressed the need for a practical tool (e.g., a leaflet) that they could bring home to read before their next consultation. It was emphasised that both the visualisation and practical tool needed be unbiased and based on the best available evidence.

### Person-centered Care as a Need for a Desirable Decision

This theme focuses on the importance of person-centred care in terms of dealing with decisional roles, taking ownership, considering beliefs and expectations, and *seeing* the individual patient in the process of decision-making. Consideration of the decisional roles, or hierarchy, within a clinical consultation, was discussed by most participants. It was evident that most of the participants recollected at least one paternalistic interaction with a healthcare practitioner, which for some was expected and accepted (**ID9**) and for others it was not accepted (**ID6**).

> **ID9**: “*I expect that the doctor will take the lead on the decision. Afterall, he is the expert. I would be uncomfortable if I was giving the entire responsibility in terms of choosing the right fit for me. I mean – I don’t know which options exists and how effective they are for my condition – He knows*”.
>
> **ID6:** “*I got the feeling, that the doctor didn’t care with me. As I said, I was much in doubt and reading through the internet and listening to everyone around me gave me more doubt. When I met my first doctor, he didn’t really listen to me worries and basically, overruled me and said I just needed to see a physio for some exercises…before I was able to say yes, he was out of the door – I just felt like the next in line*”.

Most participants expressed that decisional roles were not often addressed and discussed in the consultation, which led to decisional regrets and a feeling that the decision was ‘out of their hands’.

> **ID11**: “*There was a huge discrepancy between who needed to decide what in terms of who I spoke with while I was waiting to get my diagnosis. My general practitioner was really good at including me and asking about my preferences. However, when I ended up at the hospital, the surgeon took the decision entirely above my head. Today, I have regretted that I never spoke up – because the surgery just gave me more pain”*.

Furthermore, beliefs and expectations were considered by interview participants to be important to address in the decision-making process, to better tailor treatment to the individual. Here it was evident that the majority of participants had an expectation or belief about certain treatment options, illustrated through examples such as “*I don’t need some medical liquid (injection) in my body (**ID1**)”, “I just needed a surgery to clean up the inflammation in my shoulder* (***ID4*)**”, and “*my mother-in-law went through several months of boring exercises – I don’t want to go through that without effect (**ID13**)”*. Furthermore, the participants understood the occasionally negative influence of their beliefs and expectations in terms of choosing the right treatment and acknowledged why sometimes asking too much about this would hamper their treatment.

> **ID13**: “*looking back, my thoughts about exercise, were wrong. My physiotherapist was good at explaining to me how it worked, and she motivated me. As it turned out, I understand why clinicians sometimes must overrule our preferences”*.

Most of the participants highlighted that they needed to be actively included in the treatment decision-making, however to various extents and in specific contexts. As such, some participants explained how certain treatments facilitated more inclusion than others.

> **ID7**: “*I also believe that if I was considered for surgery, even if I wanted to, that in the end, I don’t think I should be in charge of taking the decision when the treatment is that invasive. I mean, if I had to choose between massage or exercise, that would make sense that, that was up to me”*.

Engaging PwSAPS in the decision-making process was furthermore considered important for taking ownership of their own condition, empowering them to self-manage their pain. In cases where participants articulated taking an active part in their own condition and treatment decisions, they were more motivated to adhere to their treatment plans, as evidenced by:

> **ID16**: *“The issue here is that surgical treatment is the most extreme option, so one must explore all other possibilities before resorting to surgery. I am well aware that this is necessary, which is why, this time, I have chosen to opt for self-paid physiotherapy to have a fresh perspective on the shoulder. So, physiotherapy is definitely my first choice. The second option would be a shoulder injection. I’ve had it before, and it didn’t work for me back then, so I want to try something different this time.”*

The sense of taking ownership was closely tied to also being seen as an individual person in pain. Several participants expressed that where they didn’t take ownership and engage in treatment decisions this was often accompanied by being treated like they were just on a ‘conveyor-belt’.

> **ID3**: *“So when I’m not involved, I just feel like I’m just someone who’s the next on the to-do list. I know there are guidelines, but maybe I don’t fit them. I kind of feel like that I’m more than just a guideline. I don’t feel like the treatment plan is being fitted to my situation”*.

### Supportive Environment as a Need for Adapting and Accepting the Treatment Plan

This theme describes the need for a supportive environment within which patients are readied to participate in the treatment decision-making process and are enabled to choose the right treatment for them and accept the decision. Furthermore, the theme explores how time affects the decision and how changes because of time needs to be supported. The theme also captures that emotional support is needed to help patients stay motivated, hopeful, and optimistic. PwSAPS stated that they regularly faced difficult choices, sometimes under pressure, hence requiring decisional support to effectively navigate their treatment plans. While many participants explained that they were able to support themselves in the decision-making process, several also explained that they needed the support of both their close relatives and healthcare practitioners.

> **ID14**: *“Well, the family is along for the ride. They can see when I have to step aside because I can’t be anywhere (due to pain). It’s my wife who helps lift things for me and reaches up to grab things. So, the family becomes a great support.”*

It was often noted that the certainty of decisions could change over time and that temporal changes in their condition needs to be supported throughout their healthcare journey. More specifically, some participants expressed how their life situation meant that they needed to choose different options and needed a more dynamic and adaptable healthcare plan. This was followed by participants emphasising that, through experience, they would gain new insights into their preferred pain management strategies and highlighted the need for ongoing support to re-evaluate their treatment plans.

> **ID17**: *“However, there was a point where I had to tell her (the physiotherapist) that I couldn’t handle training right now. It was because my husband had suffered an injury, and I couldn’t cope with it as he became the top priority. So, I had to take a back seat, and she fully understood. Therefore, my rehabilitation programme was put on hold for a few weeks.”*

Several participants experienced barriers to decision-making through the lack of healthcare system guidance and limited organisational resources. In this regard, it was emphasised how support in navigating the healthcare system and everything surrounding it was needed in the decision-making process. This also included access to resources, insurance, and other public services.

> **ID15**: *“I felt somewhat confused about what to do after my first consultation with the doctor, it was really quick, and I would have liked some more explanation. I think he didn’t have the time. Sometimes I don’t really think anyone have time, because it was a similar picture that I got when I met my physiotherapist. In this situation it would be helpful to have someone who can guide me, someone who isn’t a busy doctor – but I guess that don’t have the money for that”*.

Most study participants felt a need for support in dealing with the complex emotions often associated with the decision-making process. As such, it was mentioned that a trusting therapeutic alliance with their healthcare practitioner was essential to satisfactory decision-making. A trusting and respectful relationship coupled with a thorough examination were highlighted as important to help them feel included in the decision-making process and making it less stressful.

> **ID17**: *“A very, very, very pleasant guy (The doctor). He is concise and to the point, but he also takes the time to respond to what you ask. He’s not someone who talks around the issue. I like to get answers. I ask, and I ask, and I ask. So, I want to know what they do, why, and what results I can expect. And even though one may not always know it, he took the time to respond. I was very happy about that.”*

Being offered emotional support in empathic interactions with healthcare practitioners was considered important to enable good decision-making, as well as help patients adhere to the decision. Some participants explained how they had experienced various emotional challenges leading to stress and anxiety, which they believed could have been avoided if they had been given the appropriate support. Lastly, emotional support was viewed as important to enable patients to be optimistic and keep a positive outlook on their treatment plan and general life situation. Patients emphasised the importance of being surrounded by optimistic people and healthcare practitioners to reduce the decisional conflicts and regret that were present when improvements in their shoulder condition were not forthcoming.

### Mapping of Decisional Needs using Ottawa Decision Support Framework

Of the 22 ODSF decisional needs, 17 were identified based on the interviews with patient living with SAPS (see **Table 2**). The decisional needs that were not identified during the interviews were related to: social pressure, inadequate experience, inadequate instrumental help, inadequate health/social service, and inadequate financial assistance.

**Table 2.** Mapping Decisional Needs using Ottawa Decision Support Framework.

|  |  |  |
| --- | --- | --- |
| Decisional need | Code | Example of quote |
|  | Black = indicates the original code<br>Blue = indicates specific knowledge added |  |
| Decisional Need |  |  |
| - Decisional conflict | 1) Unsure about the best course of action<br>2) Worried what could go wrong about the respective treatment<br>3) Questions what is desirable<br>4) Waivers between choices | 1) ID9: “Because of my delayed diagnosis, I was kind of scared moving forward. I was so unsure about if something was broken, and because of this – I had no clue about what to do next.”<br>2) ID2: “Well, of course I trusted my doctor – but honestly, I was worried that exercise wasn’t the best fit and possibly that it could hurt my shoulder more.”<br>3) ID6: “I was much in doubt between surgery and physical therapy. I just wanted to get back to playing with my kids without pain and loss of function, but I’m not sure which option really was the best for me in the long run”.<br>4) ID9: “During my waiting time (four months), I was really struggling with whether I wanted another pair of eyes on my situation. I wouldn’t say it was |
|  |  | *on my mind every day, but I really struggled with whether I actually wanted to go with the exercise-option or if I should just seek a surgeon and get him to fix me”.* |
| - Inadequate knowledge | 1) Unaware that a decision needs to be made (e.g. person was never told they had options)<br>2) Don't know (enough) about the health problem, condition, or situation to make an informed decision<br>3) Don't know (enough) about options<br>4) Don't know (enough) about benefits, harms/risk | 1) **ID14:** “When you’re in a situation like that, you don’t think there’s a buffet of options and since you’re facing a professional who has the best option for you. So, you do what they say.”<br>2) **ID15:** “No. I had no idea. Since I hadn’t had any pain anywhere before, I thought the doctor would know. And the doctor is probably right. I accepted that I had pain in my shoulder and needed some ibuprofen.”<br>3) **ID14:** “I don’t even have that overview now. I don’t know what options are available.”<br>4) **ID14:** “Not in that way... Cons are never something that’s been mentioned. It’s funny you say that. There’s nothing I’ve thought could be a downside to, for example, doing an exercise.” |
| - Unrealistic expectations | 1) Don't know the chances of benefits, harms/risks for each option | 1) **ID13:** “I was lucky that my physiotherapist was experienced and pushed me to do exercise – otherwise, I would have no chance choosing between my options as I had only limited knowledge on the pro’s and con’s”. |
|  | 2) Perception of one's outcome probabilities are not aligned with current evidence for similar people<br>3) Difficulty believing that the outcome probabilities apply to them **and their situation** | 2) **ID3:** *"Personally, I think that if I was just being seen by a surgeon instead of a physiotherapist, I would have received surgery and I would have been much better off much quicker".*<br>3) **ID16:** *"Now I'm trying the injection. And then we'll see what happens afterwards. If it doesn't work, then I'll have to undergo surgery, personally I think that is what's best for me, but my doctor (general practitioner) doesn't agree"* |
| - Unclear values | 1) Unclear about option features that are important to them (e.g. benefits, harms/risks, other outcomes/features; scientifically uncertain outcomes) | 1) **ID11:** *"While I think my surgeon wanted the best for me, I still feel overlooked. Looking back in hindsight, if I knew how important it was to me, to somewhat fell in charge of my own health and options...then I would have done a lot thing differently".* |
| - Inadequate support and resources | 1) Inadequate perceptions **of what others think about their health**<br>2) Difficult **deciding on their** decisional | 1) **ID5:** *"My wife thought surgery was the best option to fix my shoulder quickly, but my doctor said I should try physical therapy first. Meanwhile, my friends and the internet kept telling me differently, and was all so confusing to me. At the point I didn't really knew who to listen to or what was best for me".*<br>2) **ID10:** *"I always thought I'd be the one to decide how to handle my shoulder* |
|  | roles in terms of the decision-making | *pain, but back then when the choices are in front of me, I'm unsure and went with my gut feeling. My doctor wanted to share the decision with me, but I felt sort of overwhelmed with all the medical details, but as I said I lacked more relevant information about the treatment. My family also tried to help, but at that point they were just as confused as I was. That was where I decided to trust the doctor and the physiotherapist – and yeah as I said I regretted that”.* |
|  | 3) Inadequate self-efficacy | **3) ID1:** *“As I said, I knew for sure I didn’t want an injection. But when it comes to the other options (surgery and physical therapy), I was really unsure. I didn’t trust myself to figure out which one that would have been the best for me, and honestly, I was worried that I would regret my choice.”* |
|  | 4) Inadequate motivation to make a decision | **4) ID3:** *“So when I’m not involved, I just feel like I’m just someone who’s the next on the to-do list. I know there are guidelines, but maybe I don’t fit them. I kind of feel like that I’m more than just a guideline. I don’t feel like the treatment plan is being fitted to my situation”.* |
|  | 5) Inadequate skills making a decision | **5) ID17:** *“I couldn’t answer that (choice of treatment). I was in a situation where I just needed to get home to my husband. If only he had been himself* |
|  | 6) Inadequate information available to them and their options<br>7) Inadequate advice required to make a decision<br>8) Inadequate emotional support to make a decision | *and could take care of cooking and things at home, then maybe I could have considered what should happen with me.”*<br>6) **ID5:** *“No, I wasn’t really informed about the benefits and harms of exercise. I mean that wouldn’t have been nice, as I’m not sure where I should have gotten that information elsewhere”.*<br>7) **ID2:** *“I was basically a bit tired of the clinicians not taking together – I think that would have helped me a lot – maybe my doctor would have been able to postpone my treatments if they have talked together and discussed the findings of my scan (MRI) – maybe then I could have avoided that extra pain from exercise”.*<br>8) **ID6:** *“After all this back and forth about my situation, I was sort of dealing with a lot of stress. Again, I don’t really have a lot of support”.* |
| - Complex decision characteristics | 1) Difficult decision type | 1) **ID13:** *“Deciding on treatment for my shoulder was initially almost impossible because there were so many options, and none of them guarantee a good outcome according to my doctor. She said surgery could leave me with permanent limitations, but therapy might just be a waste of time. I was* |
|  | 2) Difficult decision timing<br>3) Unreceptive to the decisional stage,<br>(e.g. because of new life-situation) | *just scared of making things worse – it felt a bit overwhelming.”*<br>2) **ID11:** *“I felt so rushed to decide on surgery for my shoulder, especially because I felt that I was overlooked throughout. As I said, I just wished that I was in other position where I confident in speaking up - I wish I had more time.”*<br>3) **ID13:** *“When I first found out about my shoulder problem, I didn’t even want to think about treatment options. I kept telling myself it wasn’t that bad, and I could just live with it. I didn’t listen to the information the doctors were giving me—I wasn’t ready to accept that I needed therapy. Looking back, I realize that I was sort of in denial, and it stopped me from really considering my options earlier.”* |
| - Personal and clinical needs | 1) Personal needs | 1) **ID6:** *“As a single mom with a full-time job, I was struggling to figure out how I’ll manage recovery time if I was going to receive surgery for my shoulder. I didn’t have the flexibility to take a lot of time off, and I don’t have a support system to help with my kids. I’m happy that it didn’t ended that way – but it might as well have ended in a disaster.”* |
|  | 2) Clinical needs | 2) **ID3:** *“I’ve been dealing with chronic pain in my shoulder for years, and it’s really taken a toll on me mentally and physically. The doctors kept giving me the same standard information about treatments, but I feel like my case is different because of how long I’ve had this issue. I feel that I need more tailored treatment that considers how this condition has affected my overall health and mental state.”* |

## Discussion

### Summary of main findings

The overall aim of this study was to explore and understand the decisional needs of patients living with SAPS. The complex nature of managing SAPS was unsurprisingly reflected in the results of the study. Participants expressed a diverse range of decisional needs during the decision-making process, including those related to their informational needs, lack of involvement, unmet expectations, unaddressed concerns, unclear values, inadequate support and resources, and uncertainty related to the influence of personal and clinical characteristics.

### Comparison with existing literature

There is a limited evidence base regarding the decisional needs of people living in pain, including shoulder pain. However, one recent Canadian study of 1649 people living with chronic pain found that most had unmet decisional needs and experienced decisional regrets during their healthcare journey [19]. Like our findings, that study found most participants preferred an active (21%, n=285) or collaborative role (76%, n=1,040) in the decision-making process [19]. Our participants indicated that active involvement in the decision-making process was often neglected, acting as a crucial influence on their compliance of treatment. This might be explained by inadequate resources for patients, where several participants mentioned that poor self-efficacy and knowledge about their condition hampered their own ability to participate actively in the decision-making process. Previous research has also highlighted that various groups experience low decisional self-efficacy and lack of information, in strong contrast to their needs [20–22]. In a recent study by Maxwell and colleagues, therapeutic alliance was identified as a key contributor to facilitating the treatment decision-making process [23]. Similarly, Malliaras et al. showed how trust in the healthcare practitioner was a key driver for patients choosing to undergo surgery [24]. These findings are supported by the results of this study, in which it was stressed that a generally supportive and trustful relationship were facilitators for a successful decision-making process.

From our study, certainty and uncertainty was frequently reported as a root for conflict, like previous research within musculoskeletal care [25–30]. The importance of addressing potential uncertainties was also highlighted by Naye et al., showing that uncertainty increased decisional conflict, emphasizing the need to address uncertainty and the underlying mechanisms causing it (e.g., lack of conditional knowledge) [19]. Our recent priority-setting study further reinforces the need for addressing uncertainties, where knowledge translation of research into clinical practice was identified as the top-priority [31]. According to the Common-Sense Model of Illness Self-Regulation, ambiguous or uncertain illness-related information can lead to patients creating cognitive and emotional representations of their illness, which guide their coping strategies and treatment decisions [32].

Both patients and healthcare providers experience inconsistency and a lack of coherence in the management of SAPS across sectors and professions [33]. Our data supports these findings in which some participants expressed large variation between healthcare practitioners in their advice and management. While previous research has primarily been focused on decision-making related to choosing a treatment [23,24], our data and previous data highlights the need for future efforts targeting decision-making across all areas of management [19]. A limiting factor for improving the overall decision-making process might be time constraints and resources in which the availability and capacity of the different professions vary substantially in clinical practice [23]. This was present in our data, where the lack of resources and organizational support was mentioned as a cause of frustration and conflicts. Barriers related to time constraints and organizational resources have been reported in the literature on multiple occasions from healthcare practitioners [34–37]. Interestingly, despite a growing body of literature showing that addressing decisional needs through SDM does not require significantly more time than practice without, it is not yet known if this applies to care of patients living with SAPS [38,39].

### Implications for clinical practice and future research

By using our findings, practitioners and future clinical aids might be more responsive to the specific concerns and challenges faced by patients living with SAPS [7,8]. As such, our findings provide guidance for clinicians wanting to address the decisional needs of the patient living with SAPS. By addressing these needs, it might be possible to reduce decisional conflicts and improve decisional outcomes. Furthermore, the findings from our study can support clinicians utilising SDM in clinical practice. Most notably, this might be achieved by providing the patients with more knowledge on their condition, their treatments options, and through active involvement in the decision-making process. The findings from this study have important implications for the design of a decision aid to support patients and clinicians in managing shoulder pain as it provides specific design features and behavioural targets for the development. To effectively support patients with SAPS in making informed decisions, future decision aids should consider incorporating simple, non-directive and unbiased information on the benefits, risks, and costs, through visual aids (e.g. charts or videos). Unbiased information is advocated by international initiatives for developing decision aids [40,41], and visuals have recently been supported as an effective means to increase patient understanding [42]. Based on our results, a future decision aid should also incorporate information on expected costs alongside treatment outcomes. A recent study highlighted that while often neglected, there may be several benefits to explicitly stating costs, including ethical aspects, reducing worries, and increasing adherence to care [43]. Furthermore, the decision aid should aim to support the patient in understanding the possibilities of taking an active role in the decision-making process. This might be aided through a series of questions or advice the patient can ask themselves or their healthcare practitioner [8,44,45]. Finally, the decision aid should be developed with input from both patients and clinicians to ensure that it meets the practical needs of clinical care and supports patients through the often-complex decision-making process. User-centered involvement is commonly used to ensure that the decision aid better addresses real-world challenges users face and is recommended by the leading guidelines on developing decision aids [47,48].

### Strengths and Limitations

To increase the validity and reliability of our findings we used two researchers for every process to prevent individual bias and subjectivity. Furthermore, through triangulation of the analysis, a more comprehensive and nuanced interpretation of the data was obtained. Another strength was the use of an empirically informed and accepted framework for establishing decisional needs. This was further strengthened by the thematic text analysis that allowed more methodological depth, which added more nuance to the decisional needs. Further, we recruited potential participants using three different methods to enhance the diversity of participants. A limitation of our study is the lack of inclusion of marginalised groups (including ethnical minorities in Denmark). A second limitation was the limited number of patients with mild to moderate SAPS (e.g., 64% of participants had consulted with surgeons). These limitations may hamper the generalisability of the results to a wider population of patients. Future studies should consider more purposive sampling to ensure a more diverse sample and utilise a multi-linguistic approach to data collection. Given our approach, we are unable to account for recall bias. Future studies should consider addressing decisional needs immediately after a consultation to avoid recall bias. Lastly, our study lacked an assessment of decisional needs through multiple perspectives, e.g. relatives and healthcare providers. Future research should consider the impact of identifying decisional needs through different stakeholders.

We explored the decisional needs for patients with SAPS. Through the ODSF, we identified 16 decisional needs for reducing decisional conflicts and regrets, and furthermore, support clinicians in addressing these needs in clinical practice. Our thematic analysis further demonstrated how patients with SAPS need certainty and information to effectively engage in the decision-making process with health-care providers. Person-centered care was deemed crucial for the participants in maximising their satisfaction with the decision-making process. Lastly, a supportive environment was necessary to support people to decide and accept treatment plans. Findings from our study indicate the need for practical tools to support patients and clinicians in improving the decision-making process in patients living with SAPS.

## Ethical approval

This study was deemed exempt from research ethical approval by the local ethics committee (ID). The study was performed in accordance with the Helsinki Declaration.

## Author contributions

All authors participated in the conceptualization, analysis, and interpretation of data, drafting, or revising the manuscript. SCB and KDL collected data. SCB, KDL, JLO, and MSR obtained funding for the study.

## Acknowledgments section

None.

## Statements and Declarations section

### Ethical considerations

The study was conducted in accordance with the Helsinki Declaration and was deemed exempt from full ethical approval by The North Denmark Region Committee on Health Research Ethics (2023-000206).

### Consent to participate

All participants gave informed consent before participation.

### Consent for publication

Not applicable.

### Data availability

Due to the nature of the research supporting data is not available to be publicly.

