## Supplemental material 1-3 for "Exploring the Decisional Needs of Patients living with Subacromial Pain Syndrome: a qualitative needs assessment study"

**Supplementary Material 1: Ottawa Decision Support Framework**

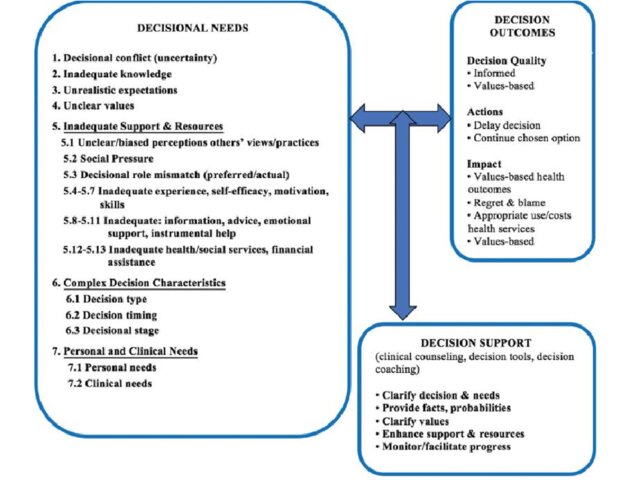

**Supplementary Material 2: Ottawa Decision Support Framework (ODSF) Decisional Needs Coding Manual**

### Conceptual/Operational Definitions

Hoefel L, O’Connor A, Lewis KB, Boland L, Sikora L, Hu J, Stacey D. (2020). 20^th^ Anniversary Update of the Ottawa Decision Support Framework part I: A systematic review of the decisional needs of people making health or social decisions. Medical Decision Making, 40(5), 555-81. Appendix B [https://doi.org/10.1177/0272989X20936209](https://doi.org/10.1177%2F0272989X20936209)

| **0.Decisional Need:** A decisional need is a deficit that can adversely affect the quality of a decision (informed, matches most valued features) and requires tailored decision support. Each need has a conceptual definition followed by an operational definition. The operational definition describes possible behavioural manifestations of the decisional need (and quantitative measures if applicable). **NEW** behavioural manifestations from the current study are **bolded**.  The codes for 22 ODSF decisional needs are shown below. To improve understanding, 18 of them are grouped under three underlined headings. | |
| --- | --- |
| 1. Decisional conflict, 2. Inadequate knowledge, 3. Unrealistic expectations 4. Unclear values 5. Inadequate support/resources (13 needs)    5.1 Inadequate perceptions: others’ views/practices    5.2 Social pressure    5.3 Difficult decisional roles    5.4 Inadequate experience    5.5 Inadequate self-efficacy    5.6 Inadequate motivation    5.7 Inadequate skills    5.8 Inadequate information    5.9 Inadequate advice    5.10 Inadequate emotional support    5.11 Inadequate instrumental help    5.12 Inadequate health/social service    5.13 Inadequate financial assistance | 6. Complex decision characteristics (3 needs)    6.1 Difficult decision type    6.2 Difficult decision timing    6.3 Unreceptive decisional stage 7. Personal and clinical needs (2 needs)    7.1 Personal needs    7.2 Clinical needs |
| 1. **Decisional Conflict** A state of personal uncertainty about which course of action to take when choice among options involve risk, loss, regret, or challenge to one’s personal values.   The hallmark manifestation of decisional conflict is verbalized uncertainty   1.1 unsure about what to do/choose/best course of action (Uncertainty subscale or item: DCS, SURE ^a.b^) Other manifestations experienced during the decision-making process or while attempting the decision**:** 1.2 worried what could go wrong; -concerned about undesired outcomes when attempting decision 1.3 wants to delay the decision 1.4 questions what is desirable /important to them (personal values) when attempting decision 1.5 feels distressed or upset when attempting decision 1.6 wavers between choices or changing one’s mind 1.7 constantly thinks about decision/can’t get off mind 1.8 feels physically stressed: tense muscles, racing heartbeat, difficulty sleeping when attempting decision | |
| 2. **Inadequate Knowledge** Unaware or lacks cognizance of essential relevant facts to make a decision: health problem/condition/situation; options; features of options (benefits, harms/risks, other outcomes/features; scientifically uncertain outcomes). Manifestations may include:    2.1 unaware that a decision needs to be made (e.g. person never told they had options)    2.2 don’t know (enough) about the health problem, condition, or situation to make a decision    2.3 don’t know (enough about) options    2.4 don’t know (enough about) benefits, harms/risks, (pros/cons, other features of options), and/or scientific uncertainties (includes medical-based outcomes and lifestyle outcomes)   (Uninformed subscale or item: DCS, SURE ^a.b^; Knowledge test: % incorrect) | |

3. **Unrealistic Expectations** Unaware of one’s chances or probabilities of outcomes (e.g. benefits, harms, other outcomes) for each option, or, perceptions of one’s outcome probabilities are not aligned with the evidence for similar people. Manifestations may include:
   3.1 don’t know chances of benefits, harms/risks for each option
   3.2 perceptions of one’s outcome probabilities are not aligned with current evidence for similar people

**NEW 3.3 difficult believing that the outcome probabilities apply to them**

(% Unrealistic expectations or inaccurate risk perceptions)

4. **Unclear Values** Lacks clarity regarding desirability or personal importance of the features of options (e.g. benefits, harms/risks, other outcomes or features; scientifically uncertain outcomes). Manifestations may include:
   4.1 unclear about option features that are important to them (e.g. benefits, harms/risks, other outcomes/features; scientifically uncertain outcomes))

(Unclear values subscale or item: DCS, SURE ^a.b^)

5. **Inadequate Support and Resources**: A grouping of 13 decisional needs related to the assistance and assets needed to make and implement the decision. They are inadequate when a person lacks the quality, appropriate quantity, and/or timely access to them. The hallmark manifestation of inadequate support and resources is:
   5.0 unsupported/lacks enough support in decision making (Unsupported subscale or item: DCS, SURE ^a.b^). The 13 individual decisional needs are:
   5.1 **Inadequate perceptions: others’ views/practices**: Perceptions of other’s views/practices refers to a person’s awareness and interpretation of what important others think is the appropriate choice (e.g. spouse, family, peers, health professional(s)). Perceptions are inadequate when a person is unaware of, lacks clarity about, or misperceives other’s views/practices or receives conflicting recommendations from others. Manifestations may include:
       5.1.1 don’t know the views/practices of others involved in the decision
       5.1.2 unclear about others’ views/practices involved in the decision
       5.1.3 misperceives others’ views/practices involved in the decision
       5.1.4 reports receiving conflicting recommendations from others
   5.2 **Social Pressure** Perception of persuasion, influence, coercion from important others (e.g. spouse, family, health professionals, or society) to choose a specific option. Manifestations may include:
       5.2.1 feels pressure from others involved in the decision (e.g. spouse, family, health professionals, or society) to choose a specific option

**5.3 Difficult Decisional Roles** Problems with one’s preferred/actual involvement in decision making. Preferred/Actual decisional roles ^c^ are classified as follows:

a) Shared: prefers to/share(s) the decision with practitioner or other, specify
b) Patient-led: prefers to/make(s) decision on their own: i) after considering others’ views, specify	, or ii) without considering others’ views
c) Delegated: prefers to/practitioner or other make(s) decision for them: i) after considering patient’s views, or ii) without considering patient’s views

Manifestations of difficult decisional roles may include:

5.3.1 Unclear about role in decision making
5.3.2 Mismatch between an informed person’s preferred role and actual role in decision making. Note, preferred roles shift when a person is informed about the nature of the decision (e.g. no clear best answer, best choice depends on what matters most to an informed person)

**NEW 5.3.3 Difficulty involving family member in decision-making. Contributing Factors may include: person did not want to worry family, family lacked knowledge**

**NEW 5.3.4 Difficult shared family deliberation on options. Contributing factors may include: different information needs, different values, communication barriers**

**NEW 5.3.5 Difficulty deliberating with practitioner because the patient/family: a) have not established a relationship with practitioner, or b) do not perceive they have positive relationship with the practitioner (e.g. trust, mutual respect, empathy, compassion, honesty, clear communication)**

**5.4 Inadequate experience** Lacks previous exposure to the condition/situation, options, outcomes and/or the decision-making process or previous experience has deleterious effects on current decision making.

Decisional experience is classified as follows: a) no previous experience; b) previous experience (for each type, specify whether positive, negative mixed): i) condition/situation; ii) options; c) outcomes; d) decision making process.

Manifestations of difficult inadequate experience may include:

5.4.1 lacks experience with
5.4.2 previous experiences had deleterious effect on current decision making, specify

5.5 **Inadequate self-efficacy** Lacks belief or confidence in one's abilities to make/implement the decision. Manifestations may include:
    5.5.1 lacks belief/confidence in ability to participate in decision-making
    5.5.2 lacks belief/confidence in ability to implement chosen option (Decision Self-Efficacy Scale)
5.6 **Inadequate motivation** Lacks desire or willingness to engage in decision making. Manifestations may include:
    5.6.1 lacks motivation or interest in making a decision
5.7 **Inadequate skills** Lacks abilities in making and implementing a decision. Manifestations may include:
    5.7.1 lacks the ability or skill to make a decision
    5.7.2 lacks the ability or skill to implement a decision
5.8 **Inadequate information** Lacks access to the quality and quantity of information (written, verbal) that is required to make and implement the decision. Manifestations may include:
    5.8.1 Lacks access to (did not receive) information about:
          5.8.1.1 to 5.8.1.7 health condition, options, benefits, harms/risks, scientific uncertainties regarding outcomes, other outcomes, other features of options
          5.8.1.8 the chances of benefits and harms/how likely each pro/con,
          5.8.1.9 what others decide or recommend,

other, specify	did not receive information **materials**

**NEW** 5.8.1.10 **other’s experiences with options (procedures, side effects, outcomes)**

5.8.2 poor quality information

**NEW 5.8.3 too much information: “information overload”**

5.9 **Inadequate advice** Lacks quality/quantity of advice required to make and implement the decision. Manifestations may include:
    5.9.1 lacks advice from important others involved in the decision
    5.9.2 poor quality advice from important others involved in the decision
5.10 **Inadequate emotional support** Lacks emotional support to make and implement the decision. Manifestations may include:
     5.10.1 lacks emotional support, specify
5.11 **Inadequate instrumental help** Lacks instrumental help to make and implement the decision. Manifestations may include:
     5.11.1 lacks instrumental help (e.g. transportation, housekeeping, daycare), specify
5.12 **Inadequate health and social services** required to make and implement the decision. Manifestations may include:
     5.12.1 lacks health & social services, specify
5.13 **Inadequate financial assistance** Lacks financial assistance to make and implement the decision. Manifestations may include:
     5.13.1 lacks financial assistance, specify

6. **Complex Decision Characteristics** A grouping of 3 decisional needs related to the features of the decision, its timing, and a person’s current phase of decision making that makes the decision more difficult.
   6.1 **Difficult Decision Type** Refers to the features of a decision that makes decision making more difficult. Decision type is classified as: a) Screening/diagnostic, b) Treatment, c) Palliative, d) Location of care, e) Other, specify	.

Features of difficult decision types may include:

6.1.1 Multiple options: n=x
6.1.2 Unknown outcomes, specify
6.1.3 Known outcomes: 6.1.3.1 to 6.1.3.5 serious effects, permanent effects (irrevocable), high chance of undesirable outcomes, outcomes valued differently by affected population, other), specify	Other decisional needs affected by difficult decisions, specify decisional need, measure (e.g. number of manifestations of DC, DCS total scale or subscales ^a^, other needs measures):
6.2 **Difficult Decision Timing** Features of the time frame for deliberation that makes decision making more difficult. Features of difficult decision types may include:
    6.2.1 Timing is urgent
    6.2.2 Decision needs to be made soon
    6.2.3 Timing is delayed

#### NEW 6.2.4 Timing is unpredictable

Other decisional needs affected by difficult decisions, specify decisional need (e.g. unreceptive decisional stage), measure (e.g. number of manifestations of DC, DCS ^a^ total scale or subscales, other needs measures):

6.3 **Unreceptive Decisional Stage** refers to the current phase of decision making: not thinking about options; actively thinking about options; close to making a choice; and taking steps or already implemented the chosen option. (Deciding not to change or to do nothing may be a viable option).

Unreceptive decisional stage is lack of openness to receive information and/or to deliberate about options in their current stage of decision making. Contributing factors may include: denial, hasty decision making, premature closure, or polarized leaning.

Decisional Stage in extracted studies are classified as: a) in process of making a decision, b) retrospectively thinking about a decision in the past, a or b) participants are either in the process of making a decision or retrospectively thinking about a decision in the past, and c) retrospectively thinking about a decision in the past and reporting on current needs now after they have made the decision

Manifestations of unreceptive decisional stage may include:

6.3.1 Unreceptive to information/deliberation, specify contributing factors:

6.3.1 to 6.3.4 denial, hasty decision making, premature closure, polarized leaning):	Other, specify:

#### NEW 6.3.5 Lack of acceptance of condition or need for treatment (from 6.3.5.1 powerful emotions,

**6.3.5.2 lack of clinical symptoms, 6.3.5.3 clinical condition (e.g. Bipolar disorder)) NEW 6.3.6 Powerful emotions affect information processing**

#### NEW 6.3.7 Unmotivated because decision too far off in the future or unpredictable

Other decisional needs affected by unreceptive decisional stage, specify decisional need, measure (e.g. number of manifestations of decisional conflict, DCS ^a^ total scale or subscales, other needs measures):

Copyright 2019 O’Connor, A.M., Hoefel, L., Lewis, K.B., & Stacey, D

**Other Needs not mapped on to ODSF,** specify

7. **Personal and Clinical Needs** A class of needs pertaining to a persons’ individual and health characteristic that can adversely affect decision quality and require decision support tailored to these special characteristics
   7.1 **Personal Needs** Special individual characteristics that can adversely affect the quality of decisions and require tailored decision support. Patient’s personal characteristics include: age, developmental stage, gender, education, marital status, ethnicity, socioeconomic status, occupation, locale. Clinical characteristics include: diagnosis & duration of condition, health status (physical, emotional, cognitive, social functioning. Practitioner characteristics are classified as follows: age, gender, ethnicity, clinical education, specialty, practice locale, experience, style of communication.

Manifestations of special personal characteristics may include:

7.1.1 Special needs due to patient’s personal characteristics, specify need/characteristic

**NEW religion/spirituality**

7.1.2 Need for tailored decision support (e.g. information or other support & resources) according to patients’ characteristics, specify need/characteristic
7.2 **Clinical Needs**

Special clinical characteristics that can adversely affect the quality of decisions and require tailored decision support. Patient’s clinical characteristics include: diagnosis & duration of condition, health status (physical, emotional, cognitive, social functioning.

Manifestations of clinical needs may include:

7.2.1 Special needs due to patient’s clinical characteristics, specify need/characteristic
7.2.2 Need for tailored decision support (e.g. information or other support & resources) according to patients’ clinical characteristics, specify need/characteristic

**Supplementary Material 3: Interview guide**

| **Subject** | **Semi-structured interview guide** | **Follow-up questions** |
| --- | --- | --- |
| **Presentation** | - Introduction, welcome - Presentation and roles - Purpose of interview (understanding the decisional needs regards to treatment options for SAPS) |  |
| **Consent** | - Confidentiality - Consent |  |
| **Background** | | |
| Demographics | - Age - Education - Treatments received |  |
| Understanding of diagnosis | - Did you receive a diagnosis at the first visit? - What explanations have the healthcare professionals you've met given you? - Which explanatory model do you think best explains your pain? - How have the explanations affected your perception of how it should be treated? | - Have you received more than one? - What has the explanation changed? |
| **Decisional Needs** | | |
| Treatment option | In regard to the first day you were examined for your pain and received a diagnosis or received an explanation for your pain:   - What were you presented with in terms of treatment options? - What familiarity did you have with different treatment options? - Throughout the process, what information have you received from healthcare professionals, online sources, or other patients regarding treatment choices? - How do you prefer to receive information about treatment choices? - What advantages and disadvantages did you think there were to the treatments you were presented with? - What advantages and disadvantages were you presented with regarding the treatment options available? | - How were you informed? By whom? - What were you told about your situation? - What were you thinking and feeling? - Do you know where and when you can receive the respective treatments? - Do you know how much time you need to allocate for the respective treatments? |
| Values and preferences | - What were your own preferences for treatment choice? - What priorities did you have regarding your choice of treatment? - What concerns did you have regarding your choice of treatment? - What expectations did you have regarding your choice of treatment? |  |
| Decision aid | - Can you tell me how the decision about which treatment to pursue was made? - Did you want to participate in the decision-making process? - How much did you want to be involved in the process yourself, and did you want others to be involved in the process as well (e.g., family members)? - When you think about the process of making a treatment choice, is there anything that could be improved? - What do you think could have prevented the above? - Were your needs for information, support, and guidance regarding the choice of treatment for your shoulder pain met? | - Were you involved in the decision about the treatment? How? - Were there others involved in the process? Who? - What were your thoughts and feelings about your treatment options? - Did you have the opportunity to ask and have questions answered? - Was there any information you lacked but did not receive? - Should the information be more/less detailed? - Should there have been more/fewer people involved in the treatment choice? - What was missing? |
| Uncertainty | - What uncertainties/concerns did you experience regarding the choice of your treatment? - How did these uncertainties/concerns affect the decision-making process? - What could be done to make you happier with the treatment you received? - In case of uncertainties/concerns, what emotions did you experience? - In case of uncertainties/concerns, what made the decision difficult? - What could make the decision easier? - What made the decision difficult, if so? - What could be done to make you happier with the treatment you received? | - How were you emotionally affected? - **Did you feel:** - Uncertainty about what to do? - Concern about what could go wrong? - Disturbed or upset? - Constantly thinking about the decision? - Wavering between choices or changing your mind? - Procrastinating the decision? - Questioning what is important to you? - Experiencing physical stress? |
